# Salience Network Segregation Mediates the Effect of Tau Pathology on Mild Behavioral Impairment

**DOI:** 10.1101/2024.05.26.24307943

**Authors:** Alexandru D. Iordan, Robert Ploutz-Snyder, Bidisha Ghosh, Annalise Rahman-Filipiak, Robert Koeppe, Scott Peltier, Bruno Giordani, Roger L. Albin, Benjamin M. Hampstead

**Affiliations:** Research Program on Cognition and Neuromodulation Based Interventions (RP-CNBI), Department of Psychiatry, University of Michigan, 4251 Plymouth Rd., Ann Arbor, MI, 48105, USA; Applied Biostatistics Laboratory, School of Nursing, University of Michigan, 426 N Ingalls St, Ann Arbor, MI 48109, USA; Department of Radiology, University of Michigan, 1500 E Medical Center Dr, Ann Arbor, MI 48109, USA; Functional MRI Laboratory, University of Michigan, 2360 Bonisteel Blvd, Ann Arbor, MI 48109, USA; Department of Biomedical Engineering, University of Michigan, 2200 Bonisteel Blvd, Ann Arbor, MI 48109, USA; Department of Neurology, University of Michigan, 1500 E Medical Center Dr, Ann Arbor, MI 48109, USA; Neurology Service & GRECC, VAAAHS, 2215 Fuller Rd, Ann Arbor, MI 48105, USA; VA Ann Arbor Healthcare System, Neuropsychology Section, Mental Health Service, 2215 Fuller Rd, Ann Arbor, MI 48105, USA

**Author notes:** **Corresponding Author**: Alexandru D. Iordan, Ph.D. University of Michigan Research Program on Cognition and Neuromodulation Based Interventions (RP-CNBI) 4251 Plymouth Rd. Ann Arbor, MI 48105.

**Keywords:** neuropsychiatric symptoms, biomarkers, positron emission tomography (PET), functional magnetic resonance imaging (fMRI), resting state, network analysis, brain connectivity

## Abstract

**INTRODUCTION:** A recently developed mild behavioral impairment (MBI) diagnostic framework standardizes the early characterization of neuropsychiatric symptoms in older adults. However, the links between MBI, brain function, and Alzheimer’s disease (AD) biomarkers are unclear.

**METHODS:** Using data from 128 participants with diagnosis of amnestic mild cognitive impairment and mild dementia – Alzheimer’s type, we test a novel model assessing direct relationships between AD biomarker status and MBI symptoms, as well as mediated effects through segregation of the salience and default-mode networks.

**RESULTS:** We identified a mediated effect of tau positivity on MBI through functional segregation of the salience network from the other high-level, association networks. There were no direct effects of AD biomarkers status on MBI.

**DISCUSSION:** Our findings suggest an indirect role of tau pathology in MBI through brain network dysfunction and emphasize the role of the salience network in mediating relationships between neuropathological changes and behavioral manifestations.

## 1. BACKGROUND

Later-life neuropsychiatric symptoms are increasingly acknowledged as additional early symptoms of Alzheimer’s disease (AD) and related dementias (ADRD), either independently of, or alongside mild cognitive impairment (MCI).^1–5^ Over half of the individuals diagnosed with cognitive disorders, including dementia, develop neuropsychiatric symptoms before onset of cognitive impairments, often several years earlier.^4^ A recently proposed mild behavioral impairment (MBI) diagnostic framework standardizes assessment of neuropsychiatric symptoms in older adults.^6, 7^ The neurophysiological mechanisms of MBI within AD/ADRD are relatively understudied, which results in an incomplete understanding and limits effective treatment options.^8^

Earlier studies reported MBI was associated with accumulation of AD pathology and medial temporal lobe atrophy,^9–11^ suggesting that early neuropathological changes of AD play key roles in MBI. However, emerging evidence linking MBI to widespread structural and functional brain changes, including in frontal and parietal regions,^12–16^ suggests that focusing solely on the temporal lobe is an overly simplistic approach, insufficient for understanding complex, network-level dysfunctions in MBI. Network-based approaches align with the established role of the brain’s functional network architecture in the progression of AD pathologies^17^ and provide an opportunity to better understand associations between AD pathologies and their clinical manifestations. Previous studies employing resting-state connectivity identified AD-related disruptions in functional connectivity within and between the major brain networks, including the salience and default-mode networks.^17–20^ The salience network is anchored in the anterior insula, dorsal anterior cingulate and supramarginal cortices, and is associated with the detection of, and initial response to, external inputs and internal events with emotional or motivational value.^21–23^ The default-mode network is anchored in the medial prefrontal, posterior cingulate and lateral inferior parietal cortices, and is associated with memory and self-referential processing,^24^ the integration of cognitive and emotional experience, and the regulation of emotional responses.^25–27^ Functional connectivity disruptions of the salience and default-mode networks are linked to neuropsychiatric symptoms along the dementia spectrum.^12, 28–31^ Recent functional magnetic resonance (fMRI) evidence, based on a mixed sample of dementia-free individuals with or without MCI, points to lower functional connectivity within the salience and default-mode networks for individuals with MBI compared to those without MBI.^15^ To our knowledge, we are the first to ask how AD pathologies relate to salience and default-mode network dysfunctions and whether such relationships are associated with MBI severity. It is also unclear what role the frontoparietal network plays in the relationship between clinical features and AD pathology, given its role in cognitive control^32^ and the potential dysfunction of this network in those with MBI.^12^

To address this gap in the literature, we focus on the relationships between (1) AD pathology measured using amyloid-beta (Aβ) and tau positron emission tomography (PET); (2) brain network segregation—an index of functional network integrity—defined as the balance of within– and between-network connectivity strength,^33–35^ measured during resting-state fMRI; and (3) MBI symptoms indexed using the Neuropsychiatric Inventory Questionnaire (NPI-Q), a widely-used informant-rated instrument for the assessment of neuropsychiatric symptoms.^36^ Using path analysis, we test a novel model comprising direct relationships between AD biomarker status (i.e., AD biomarker negative, Aβ positive and tau negative, and both Aβ and tau positive) and MBI symptoms, as well as mediated effects through the segregation of the salience and default-mode networks. Based on recent evidence,^15^ we hypothesize that network segregation of the salience and default-mode networks mediates the effect of AD pathology on MBI severity.^26, 27, 37^ As a control analysis, we also test an alternative model comprising the frontoparietal control network, in addition to the salience and default-mode networks, to evaluate potential contributions of the frontoparietal control network dysfunction to MBI.^12^

## 2. METHODS

### 2.1. Participants

We evaluated data from 136 participants enrolled in the “Stimulation to Improve Memory” (STIM) study (R01AG058724; NCT03875326; PI Hampstead) who completed neuropsychological, structural and functional MRI, and Aβ and tau PET assessments. The STIM study included participants aged 55 or older, with a diagnosis of amnestic MCI or mild dementia of the Alzheimer’s type (DAT), who were MRI compatible (using the American College of Radiology guidelines)^38^ and stable on relevant medications for at least 4 weeks prior to study enrollment. Participants were excluded if they had other neurological disease (e.g., epilepsy, stroke, traumatic brain injury), psychiatric conditions (e.g., moderate-severe depression, bipolar disorder, schizophrenia), sensory impairments that limited the ability to take part in the study, or significant history of, or current alcohol or drug abuse/dependence. Eight participants were excluded due T1-weighted anatomical image segmentation error (5 participants) or excessive in-scanner motion (3 participants; see *MRI data acquisition and processing* below). Thus, our final sample included 128 participants. Demographic characteristics are presented in Table 1. The University of Michigan Institutional Research Board approved this study. All participants or their legally authorized representative provided written informed consent (and assent as appropriate).

**Table 1.**
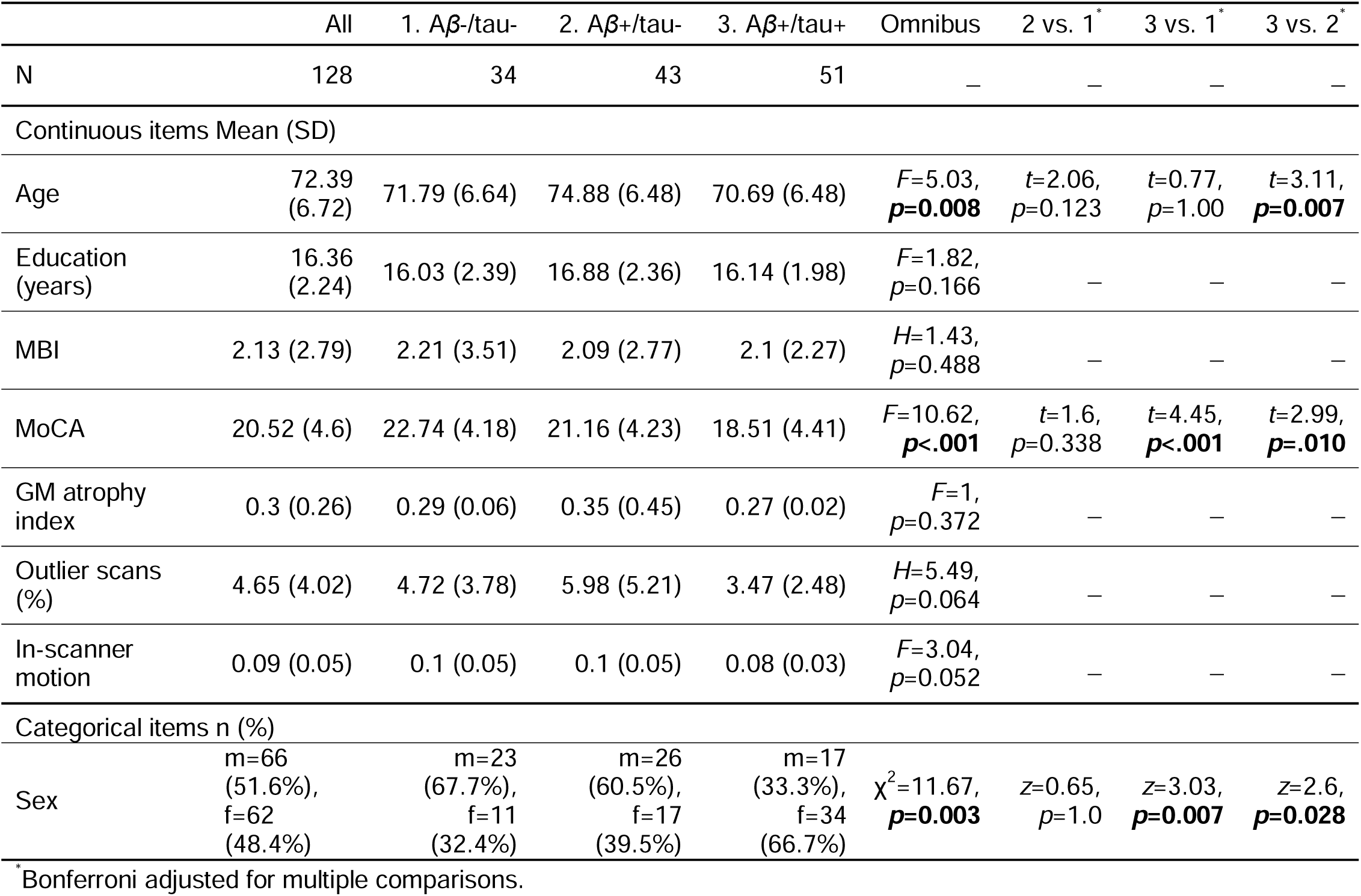
Demographic, neuropsychological, and MRI characteristics of our sample. Abbreviations: A*β*-/tau-: AD biomarker negative; A*β*+/tau-: A*β* positive/tau negative; A*β*+/tau+: A*β* positive/tau positive; m: male; f: female; MBI: mild behavioral impairment; MoCA: Montreal Cognitive Assessment; GM: grey matter.

### 2.2. Measures

Demographic characteristics (age, sex and education) and information regarding medical comorbidities (based on diagnoses by physicians) were collected as part of participants’ overall medical history and supplemented with data from participants, caregivers, and medical records. Participants underwent comprehensive neuropsychological evaluation using the Uniform Data Set (v3.0) and additional measures, including the Montreal Cognitive Assessment (MoCA)^39^, to assess cognitive function and the informant-rated Neuropsychiatric Inventory Questionnaire (NPI-Q),^36^ completed by a close informant, to assess neuropsychiatric symptoms.

Mild Behavioral Impairment (MBI) scores were derived from the NPI-Q results using a published algorithm.^40, 41^ These scores encompassed five domains: decreased motivation (NPI-Q apathy; range 0-3), emotional dysregulation (NPI-Q depression, anxiety, and elation; range 0-9), impulse dyscontrol (NPI-Q agitation, irritability, and motor behavior; range 0-9), social inappropriateness (NPI-Q disinhibition; range 0-3); abnormal perception or thought content (NPI-Q delusions and hallucinations; range 0-6). Global MBI burden was calculated by summing these domain scores (range 0-30).^42–44^ Of note, the reference range for NPI-Q is 1 month, and thus the scoring presumably reflected the immediately prior one-month period.^41^

### 2.3. MRI data acquisition and processing

Imaging data were collected using a 3T General Electric Discovery scanner with a 32-channel Nova Medical head coil. Resting-state data were acquired in interleaved ascending order using a gradient echo sequence, with MR parameters: TR/TE=800/30ms; multiband factor=6; flip angle=52°; field of view=216×216mm^2^; matrix size=90×90; slice thickness=2.4mm, no gap; 60 axial slices; voxel size=2.4×2.4×2.4mm^3^. After an initial 9.6 seconds of signal stabilization, 570 volumes of resting-state were acquired. A high-resolution T1-weighted anatomical image was also collected preceding resting-state acquisition, using spoiled-gradient-recalled acquisition (SPGR) in steady-state imaging (TR/TE=6.95/2.92ms, flip angle=8°, field of view=256×256mm^2^, matrix size=256×256; slice thickness=1mm; 208 sagittal slices; voxel size=1×1×1mm^3^).

Preprocessing was performed using SPM12 (Wellcome Department of Cognitive Neurology, London) and MATLAB R2019b (The MathWorks Inc., Natick, MA). Functional images were slice-time corrected, realigned and field-map corrected, and co-registered to the anatomical images. A study-specific anatomical template was created, using Diffeomorphic Anatomical Registration Through Exponentiated Lie Algebra (DARTEL),^45^ based on segmented grey matter and white matter tissue classes, to optimize inter-participant alignment.^46^ The DARTEL flowfields and MNI transformation were then applied to the functional images, and the functional images were resampled to 3×3×3mm^3^ voxel size. The average proportion of outlier scans (differential motion *d*>0.5mm or global intensity *z*>3, identified with Artifact Detection Toolbox [ART]; nitrc.org/projects/artifact_detect/) was 4.6%. As mentioned in the *Participants* section above, 3 participants had >25% of data marked as outliers and thus were excluded from the analyses.

Brain-wide functional connectivity analyses were performed using the Connectivity Toolbox (CONN)^47^ and the commonly used Power et al.^48^ functional atlas. A 5 mm-radius sphere was centered at each atlas coordinate. We used 212 regions of interest that showed overlap with our functional data. To remove physiological and other sources of noise from the fMRI time series we used linear regression and the anatomical CompCor method.^49–51^ Each participant’s white matter and cerebrospinal fluid masks derived during segmentation, eroded by 1 voxel to minimize partial volume effects, were used as noise ROIs. The following temporal covariates were added to the model: undesired linear trend, signal extracted from each participant’s noise ROIs (5 principal component analysis parameters for each), motion parameters (3 rotation and 3 translation parameters, plus their first-order temporal derivatives), regressors for each outlier scan (i.e., “scrubbing”; one covariate was added for each outlier scan, consisting of 0’s everywhere but the outlier scan, coded as “1”). The residual fMRI time series were bandpass filtered (0.008-0.09Hz). Functional connectivity was estimated using a Pearson correlation between each pair of time series. Finally, the correlation coefficients were Fisher-*z* transformed, and the diagonal of the connectivity matrix was set to zero.

We computed the network segregation metric^34, 35^ using the Power et al.^48^ node-module assignments. Network segregation was calculated as the difference of within– and between-networks connectivity expressed as a proportion of within-network connectivity, i.e. (Z_w_-Z_b_)/Z_w_, where Z_w_ is the mean within-network connectivity and Z_b_ is the mean between-network connectivity. In line with prior investigations,^34, 48, 52^ negative connectivity values were set to zero before calculating segregation, due to continuing debates regarding their interpretation, particularly during resting-state recordings.^50, 53, 54^

Given our *a priori* hypotheses, we targeted the canonical salience, default-mode, and frontoparietal control networks (Supplementary Figure 1). These three networks are the core association networks (i.e., networks that cover “association” areas of the brain)^34^ included in the “triple-network model” of brain function^55^ and psychopathology.^18^ The remaining three association networks in the Power et al.^48^ atlas include the cingulo-opercular network, mainly related to initiating and sustaining goal-oriented behavior,^56^ and the dorsal and ventral attention networks, typically linked to attention allocation toward the external environment.^57^ For completeness, we included these latter three networks in computing the overall indices of network segregation within the association system;^34^ however, we had no specific hypotheses for their roles in the relationship between AD biomarkers and MBI. We first calculated the overall segregation of each of the targeted three networks (i.e., salience, default-mode, and frontoparietal networks, respectively) relative to all the other five association networks. Within-network connectivity was calculated for each targeted network and between-network connectivity was calculated for each targeted network relative to all the other five association networks. Then, we calculated pairwise segregation between any two targeted networks (i.e., salience–default-mode, default-mode–frontoparietal, and salience–frontoparietal, respectively), with within– and between-network connectivity calculated only relative to the two networks in each pair.

### 2.4. PET data acquisition and processing

Two PET radiotracers were used in this study, one to assess Aβ levels ([^11^C]PiB) and one to assess tau levels ([^18^F]AV-1451) in the brain. [^11^C]PiB scans were acquired as an 80-min, 23-frame dynamic sequence as follows: 4×0.5min, 3×1min, 2×2.5min; and 14×5min. A partial bolus plus constant infusion protocol was used administering 18 mCi of PiB, with 45% given as a bolus over the first minute of the study, followed by a continuous infusion of the remaining 55% of the dose over 80 minutes. [^18^F]AV1451 (flortaucipir; tauvid) scans were acquired as a 70-min, 16-frame dynamic sequence as follows: 4×0.5min, 3×1min, 2×2.5min, 2×5min, and 5×10min. A bolus plus infusion protocol was also used for [^18^F]AV1451, with 60% given as a bolus over the first minute of the study, followed by a continuous infusion of the remaining 40% of the dose over 70 minutes. The PET scans were acquired on a Siemens BioGraph TruePoint Model 1094 PET/CT system (Siemens Healthcare, Knoxville, TN). Image reconstruction was performed using 3D-OSEM, with 4 iterations, 21 subsets into a 336 matrix and a 3mm Gaussian post-reconstruction filter for both Aβ and tau radiotracers.

All dynamic PET image frames were co-registered within subjects using a rigid-body transformation to reduce the effects of subject motion during the imaging session (NeuroStat; neurostat.neuro.utah.edu). Parameter estimation for both Aβ and tau radiotracers was performed using an equilibrium approach, which is afforded by the use of the bolus+infusion protocols described above and allows brain concentrations to approach steady-state during the PET study. Distribution volume ratio (DVR) images were calculated as the late-frame image average divided by the reference-region concentration at the end of the study. For PiB, the time frames from 40-80 min and a whole-cerebellum reference region were used to estimate Aβ DVR. For AV1451, the time frames from 40-70 min and a cerebellar gray matter reference regions were used to estimate tau DVR.

Different approaches were used to extract volume of interest (VOI) measures for the Aβ and tau scans. For PiB, the NeuroStat software package was used to orient the co-registered image set into the stereotactic coordinate system of the Talairach atlas and a template of atlas VOIs were applied to the dynamic scans with regions including frontal, parietal, temporal, and occipital cortices, putamen, caudate nucleus, thalamus, hippocampus, amygdala, cerebellar and brainstem regions. In addition, the Centiloid method (CL)^58^ was used to calculate a CL value for each subject from a large cortical region and the whole-cerebellum reference region. A previous series of bolus vs. bolus+infusion PiB scans was used to calculate the ‘bolus+infusion’ SUVr to CL conversion equation based on the methods described in the original Centiloid paper.^58^

For the AV-1451 tau scans, we used FreeSurfer^59^ to register the dynamic PET sequence and DVR images to the subject’s own MR scan. FreeSurfer was used to warp the entire set of FreeSurfer atlas regions onto individual’s MR and hence their co-registered PET images. For tau VOIs, we used the same combination of FreeSurfer regions for left and right hemispheres as reported in Royse et al.^60^ to create Braak areas 1 through 6 and the Braak meta-region.

Finally, we classified as Aβ positive individuals with Aβ CL≥ 20CL^60^ and as tau positive individuals with tau SUVr≥1.25 in either left of right Braak meta-region (>4SD above our reference group mean). Using these criteria, our sample comprised three groups: (1) individuals who were AD biomarker negative (Aβ-/tau-), (2) individuals who were Aβ positive and tau negative (Aβ+/tau-), and (3) individuals who were both Aβ and tau positive (Aβ+/tau+). There were no individuals who were Aβ negative and tau positive (see Table 1).

### 2.5. Statistical analysis

Analyses were performed using Mplus^61^ and Stata^62^ statistical software. All statistical assumptions were tested prior to hypothesis testing, setting α=0.05 for statistical significance. We conducted two path analyses designed to evaluate the direct and indirect (mediated) relationships between AD biomarker status and MBI, with the latter modeled as a negative binomially distributed outcome.

Our main model focused on the direct predictive pathways between AD biomarker status and MBI, with potentially mediated pathways through the overall segregation of the salience and default-mode networks from the other association networks and pairwise salience–default-mode segregation. This model was designed to evaluate the hypothesis that the effects of AD biomarker status on MBI are at least partially mediated through brain network dysfunctions. Given the number of parameters involved in this path-analytic approach and acknowledging the relatively small sample size (n=128) relative to model complexity, we also applied a more traditional mediation analysis,^63^ which yielded comparable results (see Supplementary Analysis).

We then expanded this model (i.e., control analysis) to incorporate potential mediation pathways through the frontoparietal control network, in addition to the salience and default-mode networks, as well as the pairwise segregations between each two networks (i.e., salience–default-mode, default-mode–frontoparietal, and salience–frontoparietal). This alternative model was designed as a follow-up to the first model to evaluate the specificity of the association between AD biomarker status and MBI through the salience and default-mode networks, and not more globally through frontoparietal network function. Both models incorporated demographic characteristics (age, sex, and education), cognitive performance (assessed using MoCA), brain atrophy (ratio of cortical grey matter volume to total intracranial volume, both estimated using FreeSurfer), and neuroimaging data quality (percent outlier scans and in-scanner motion, i.e. mean displacement calculated using ART, as described above) as model covariates in order to evaluate the associations of interest while controlling for these covariates.

## 3. RESULTS

### 3.1. Demographic, neuropsychological, and MRI characteristics

Demographic, neuropsychological, and MRI characteristics of our sample are presented in Table 1. There were significant age differences across the three AD biomarker groups (*F*=5.03, *p*=0.008). Specifically, individuals who were Aβ+/tau-were older than those who were Aβ+/tau+ (*t*=3.11, *p*=0.007). In addition, the male-to-female ratio was different across the three groups (χ^2^=11.67, *p*=0.003), with the Aβ+/tau+ group including more women than the Aβ-/tau-(*z*=3.03, *p*=0.007) and Aβ+/tau-(*z*=2.6, *p*=0.028) groups. Also, there were differences in global cognition (as assessed by MoCA) across the three groups, with the Aβ+/tau+ group showing lower cognitive performance than both the Aβ-/tau-(*t*=4.45, *p*<0.001) and Aβ+/tau-(*t*=2.99, *p*=0.01) groups, which is an expected pattern of findings.

### 3.2. Main model: salience and default-mode networks

Our main model (Figure 1) included direct paths from AD biomarkers to MBI and mediated pathways through the overall segregation of the salience and default-mode networks from the other association networks, as well as the pairwise salience–default-mode segregation. Results showed a mediated path of the effect of Aβ+/tau+ biomarker status on MBI severity through the overall segregation of the salience network (indirect effect: β=0.07, *p*=0.044).

**Figure 1.**
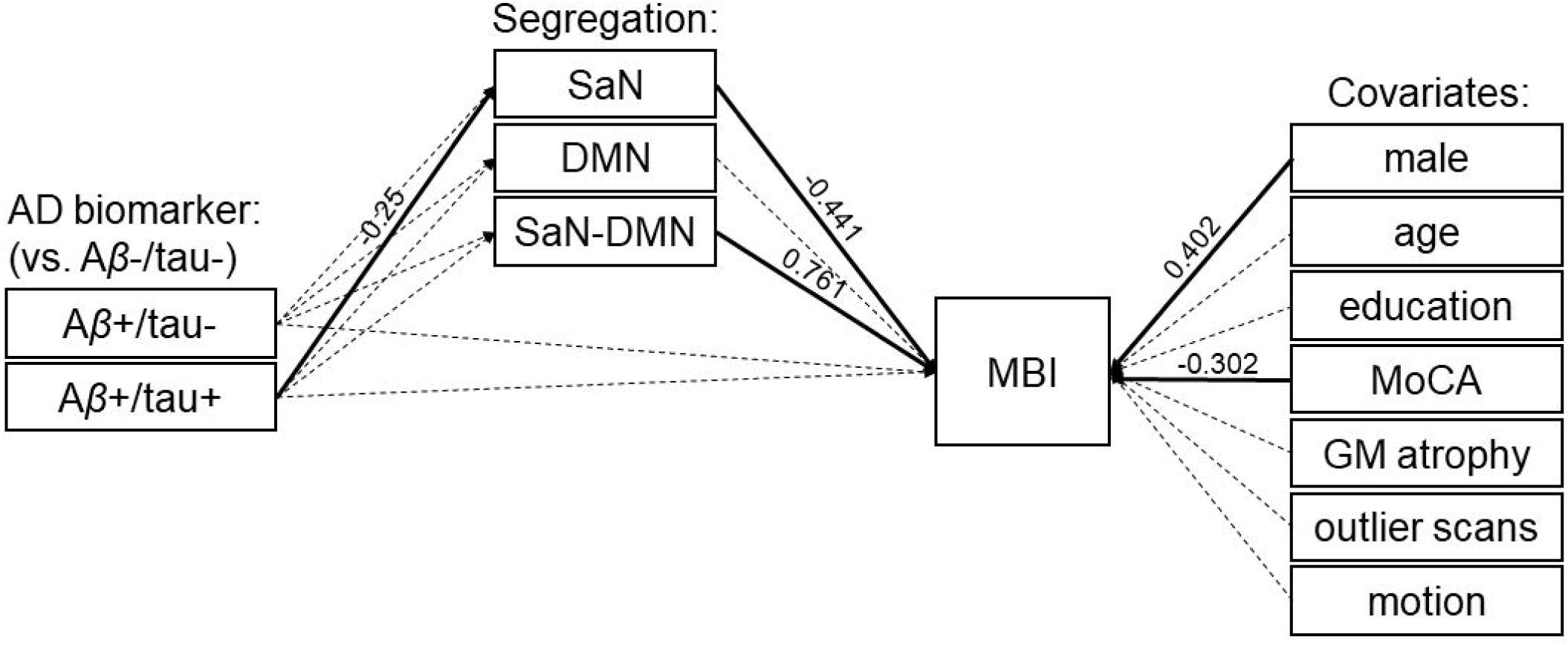
Main model. The overall segregation of the salience network (SaN) from the other association networks mediates the effect of Aβ positive/tau positive (Aβ+/tau+) biomarker status on MBI (indirect effect: β=0.07, *p*=0.044). There is no direct path from AD biomarkers to MBI. Solid and dashed arrows indicate significant and non-significant relationships, respectively (see Table 2). Abbreviations: AD: Alzheimer’s disease; Aβ-/tau-: AD biomarker negative; Aβ+/tau-: Aβ positive/tau negative; Aβ+/tau+: Aβ positive/tau positive; SaN: salience network; DMN: default-mode network; MBI: mild behavioral impairment; MoCA: Montreal Cognitive Assessment; GM: grey matter.

**Table 2.** Path analysis results for the main model. Abbreviations: MBI: mild behavioral impairment; A*β*+/tau-: A*β* positive/tau negative; A*β*+/tau+: A*β* positive/tau positive; Seg.: network segregation; SaN: salience network; DMN: default-mode network.

| Variables | Predictor | Beta | S.E. | P-Value |
| --- | --- | --- | --- | --- |
| *Outcome* |  |  |  |  |
| **MBI** |  |  |  |  |
| | $A\beta$+/tau- | -0.025 | 0.161 | 0.879 |
| | $A\beta$+/tau+ | 0.017 | 0.177 | 0.922 |
|  | Seg. SaN | -0.441 | 0.178 | **0.013** |
|  | Seg. DMN | -0.179 | 0.187 | 0.339 |
|  | Seg. SaN-DMN | 0.761 | 0.232 | **0.001** |
| *Covariates* |  |  |  |  |
|  | male | 0.402 | 0.118 | **0.001** |
|  | age | -0.027 | 0.126 | 0.831 |
|  | education | -0.139 | 0.116 | 0.229 |
|  | MoCA | -0.302 | 0.109 | **0.006** |
|  | GM atrophy | 0.041 | 0.033 | 0.209 |
|  | outlier scans | -0.164 | 0.137 | 0.229 |
|  | motion | 0.175 | 0.124 | 0.157 |
| *Mediators* |  |  |  |  |
| Seg. SaN |  |  |  |  |
| | $A\beta$+/tau- | -0.163 | 0.102 | 0.109 |
| | $A\beta$+/tau+ | -0.25 | 0.093 | **0.007** |
| Seg. DMN |  |  |  |  |
| | $A\beta$+/tau- | -0.054 | 0.112 | 0.632 |
| | $A\beta$+/tau+ | 0 | 0.1 | 0.997 |
| Seg. SaN-DMN |  |  |  |  |
| | $A\beta$+/tau- | -0.123 | 0.115 | 0.283 |
| | $A\beta$+/tau+ | -0.134 | 0.113 | 0.232 |

Specifically, relative to biomarker-negative individuals, those who were positive for both Aβ and tau showed lower overall segregation of the salience network (path from Aβ+/tau+ to salience network segregation: β=-0.25, *p*=0.007). Furthermore, individuals showing lower overall segregation of the salience network had greater severity of MBI symptoms (path from salience network segregation to MBI: β=-0.441, *p*=0.013). There were no direct paths from AD biomarker status to MBI (*p*>0.8; see Table 2). In addition, there was a significant effect of the pairwise salience–default-mode segregation on MBI symptom severity (β=0.761, *p*=0.001). Finally, men were positively associated (β=0.402, *p*=0.001) and MoCA scores were negatively associated (β=-0.302, *p*=0.006) with the severity of MBI symptoms.

### 3.3. Alternative model: salience, default-mode, and frontoparietal control networks

Our alternative model (Figure 2) expanded the main model to include potential mediation pathways through the overall segregation of the frontoparietal control network from the other association networks, as well as the pairwise segregation between the frontoparietal control network and the salience and default-mode networks, respectively. Consistent with our main model, results showed a mediated path of the effect of Aβ+/tau+ status on the severity of MBI symptoms through the overall segregation of the salience network (indirect effect: β=0.08, *p*=0.039). Specifically, relative to biomarker-negative individuals, those who were positive for both Aβ and tau showed lower overall segregation of the salience network (path from Aβ+/tau+ to salience network segregation: β=-0.25, *p*=0.007). Furthermore, individuals showing lower overall segregation of the salience network had greater severity of MBI symptoms (path from salience network segregation to MBI: β=-0.48, *p*=0.012). There were no direct paths from AD biomarker status to MBI (*p*>0.8; see Table 3). In addition, there were significant effects of Aβ+/tau+ biomarker status on the pairwise salience–frontoparietal network segregation (β=-0.301, *p*=0.005) and of the pairwise salience–default-mode network segregation on MBI symptom severity (β=0.785, *p*=0.001). Finally, men were positively associated (β=0.388, *p*=0.002) and MoCA scores were negatively associated (β=-0.309, *p*=0.005) with the severity of MBI symptoms.

**Figure 2.**
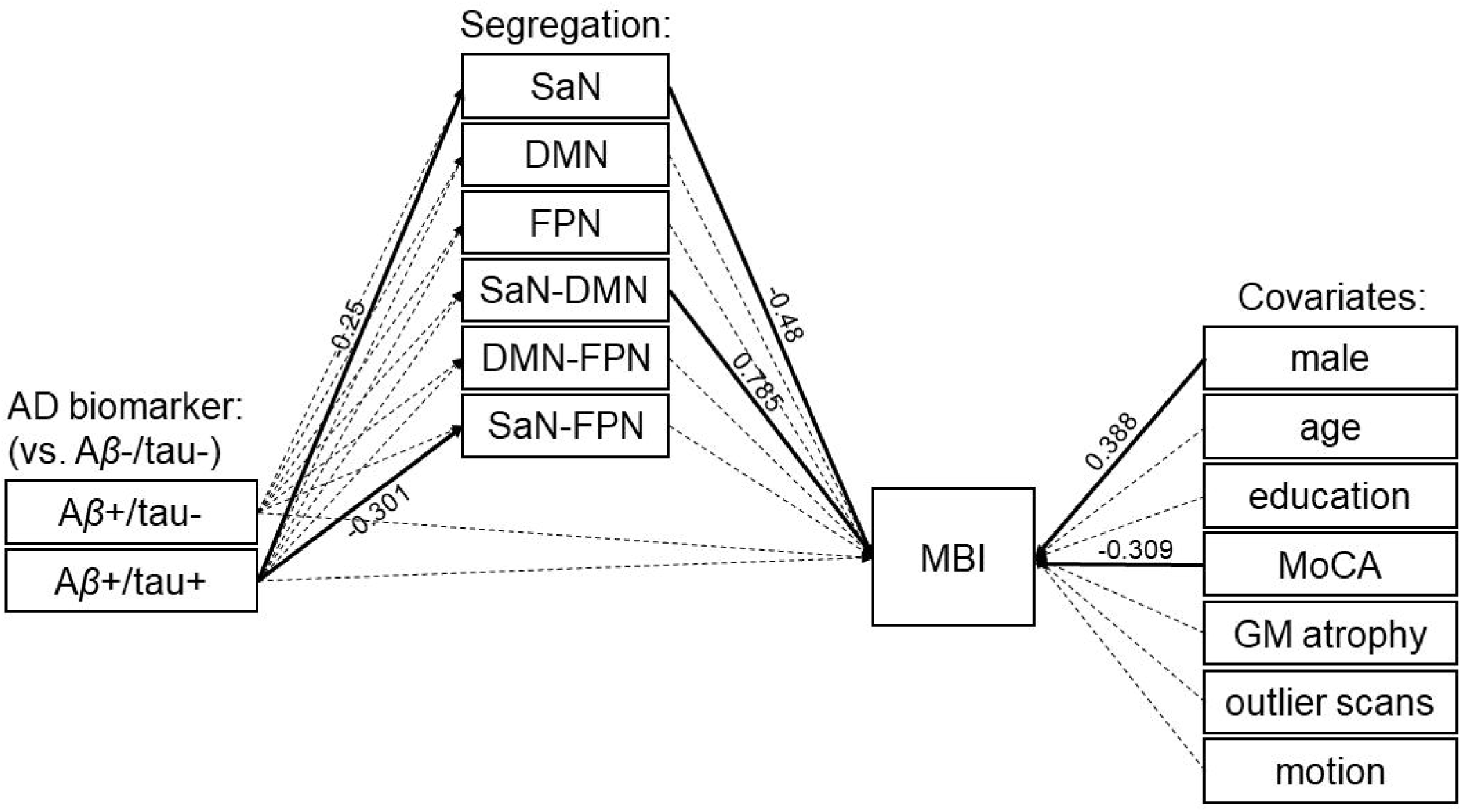
Alternative model. Similar to our main model, only the overall segregation of the salience network (SaN) from the other association networks mediates the effect of Aβ and tau positivity (Aβ+/tau+) on MBI (indirect effect: β=0.08, *p*=0.039). There is no direct path from AD biomarkers to MBI. Solid and dashed arrows indicate significant and non-significant relationships, respectively (see Table 3). Abbreviations: AD: Alzheimer’s disease; Aβ-/tau-: AD biomarker negative; Aβ+/tau-: Aβ positive/tau negative; Aβ+/tau+: Aβ positive/tau positive; SaN: salience network; DMN: default-mode network; MBI: mild behavioral impairment; MoCA: Montreal Cognitive Assessment; GM: grey matter.

**Table 3.** Path analysis results for the alternative model. Abbreviations: MBI: mild behavioral impairment; A*β*+/tau-: A*β* positive/tau negative; A*β*+/tau+: A*β* positive/tau positive; Seg.: network segregation; SaN: salience network; DMN: default-mode network; FPN: frontoparietal control network.

| Variables | Predictor | Beta | S.E. | P-Value |
| --- | --- | --- | --- | --- |
| *Outcome* |  |  |  |  |
| **MBI** |  |  |  |  |
| | $A\beta$+/tau- | -0.001 | 0.164 | 0.997 |
| | $A\beta$+/tau+ | 0.037 | 0.177 | 0.835 |
|  | Seg. SaN | -0.48 | 0.192 | **0.012** |
|  | Seg. DMN | -0.169 | 0.213 | 0.429 |
|  | Seg. FPN | -0.058 | 0.2 | 0.771 |
|  | Seg. SaN-DMN | 0.785 | 0.237 | **0.001** |
|  | Seg. SaN-FPN | 0.101 | 0.156 | 0.52 |
|  | Seg. DMN-FPN | -0.041 | 0.19 | 0.829 |
| *Covariates* |  |  |  |  |
|  | male | 0.388 | 0.123 | **0.002** |
|  | age | -0.028 | 0.13 | 0.832 |
|  | education | -0.164 | 0.116 | 0.157 |
|  | MoCA | -0.309 | 0.111 | **0.005** |
|  | GM atrophy | 0.058 | 0.042 | 0.172 |
|  | outlier scans | -0.15 | 0.141 | 0.287 |
|  | motion | 0.152 | 0.128 | 0.235 |
| *Mediators* |  |  |  |  |
| Seg. SaN |  |  |  |  |
| | $A\beta$+/tau- | -0.163 | 0.102 | 0.109 |
| | $A\beta$+/tau+ | -0.25 | 0.093 | **0.007** |
| Seg. DMN |  |  |  |  |
| | $A\beta$+/tau- | -0.054 | 0.112 | 0.632 |
| | $A\beta$+/tau+ | 0 | 0.1 | 0.997 |
| Seg. FPN |  |  |  |  |
| | $A\beta$+/tau- | -0.099 | 0.102 | 0.331 |
| | $A\beta$+/tau+ | -0.085 | 0.109 | 0.436 |
| Seg. SaN-DMN |  |  |  |  |
| | $A\beta$+/tau- | -0.123 | 0.115 | 0.283 |
| | $A\beta$+/tau+ | -0.134 | 0.113 | 0.232 |
| Seg. SaN-FPN |  |  |  |  |
| | $A\beta$+/tau- | -0.163 | 0.106 | 0.123 |
| | $A\beta$+/tau+ | -0.301 | 0.106 | **0.005** |
| Seg. DMN-FPN |  |  |  |  |
| | $A\beta$+/tau- | 0.06 | 0.103 | 0.561 |
| | $A\beta$+/tau+ | 0.065 | 0.106 | 0.537 |

Model fit: AIC = -579.663; BIC = -462.73; Sample-Size Adjusted BIC = -592.394.

## 4. DISCUSSION

We identified a mediated effect of tau positivity on MBI through functional segregation of the salience network from the other high-level, association networks. Specifically, individuals who were positive for both Aβ and tau showed lower overall segregation of the salience network compared to those who were AD biomarker negative. Lower overall segregation of the salience network was associated with more severe MBI symptoms. Taken together, these findings suggest that the effect of tau pathology on MBI may operate through the modulation of salience network functional integrity. This mediated relationship was unique for the salience network since including the default-mode and frontoparietal control networks in our models did not change the tau-specific findings. To the best of our knowledge, this is the first study to integrate MBI with AD PET biomarkers and network-level brain functionality within a unified framework.

Although initial investigations linking MBI with AD biomarkers suggested a role of Aβ pathology,^64^ subsequent findings indicated tau as a more critical factor.^9, 65^ Our study supports this latter perspective, specifically that accumulation of tau pathology may be the main driver of MBI symptomatology. Moreover, our results extend these previous findings by identifying salience network dysfunction as a potential mechanism though which tau pathology influences MBI symptoms.

In the triple-network model,^18, 55^ the salience network is thought to implement shifting between external and internal “modes” of processing via coordination with the frontoparietal control and default-mode networks, respectively. The salience network is also posited to play a major role in emotional awareness.^26^ Its major hub, the anterior insula, is proposed as a site of integration between “bottom-up” interoceptive signals (i.e., homeostatic and metabolic input from the body) with “top-down” cognitive predictions to form a representation of one’s physiological state and relay this information to subjective awareness as affective feelings.^37^

Our findings are consistent with the interpretation that disruptions in salience network functionality may drive behavioral symptoms in individuals with AD-related tau pathology. The observed *indirect* role of tau pathology in MBI through the salience network emphasizes its role in mediating the relationship between neuropathological changes and behavioral manifestations. This supports an emerging view of neurodegenerative diseases as network-level disorders, where clinical symptoms are closely tied to the functional integrity of specific brain networks.^17^ Brain network abnormalities likely act as an intermediate phenotype between neuropathological changes and clinical syndromes. This could explain complex relationships where the same primary type of pathology manifests as different clinical phenotypes, and conversely, similar clinical presentations may arise from different neuropathological origins.^17^

Our results showing reduced functional segregation of the salience network in tau-positive individuals align with documented changes in the salience network’s connectivity as AD progresses.^66^ While network segregation typically decreases with aging,^34, 67, 68^ this process is exacerbated in AD.^69, 70^ Progression of AD is marked by a nonlinear pattern of connectivity within the salience and default-mode networks, transitioning from initial *hyper*connectivity associated with Aβ accumulation and low tau levels to *hypo*connectivity as tau pathology accumulates and neuronal damage becomes more extensive.^17, 66, 69^ Our findings of lower salience network segregation in tau-positive individuals with cognitive symptoms suggest we are capturing the hypoconnectivity stage, when brain networks become less discrete and more blended in their function.

The temporal transition from salience network hyper-to hypoconnectivity with AD progression may explain previously reported associations between neuropsychiatric symptoms and salience network hyperconnectivity.^28, 71, 72^ Although neuropsychiatric symptoms in early-stage AD have been linked with enhanced salience network connectivity,^28, 71^ the nature of this relationship in the later stages of AD—characterized by a shift towards salience network hypoconnectivity—is less clear.^66, 69^ Our findings are also consistent with recent evidence based on a mixed sample of dementia-free individuals with or without MCI, showing lower functional connectivity within the salience network for individuals with MBI relative to those without MBI.^15^

Finally, analyses for the main model also showed a positive association between the pairwise salience–default-mode network segregation and the severity of MBI symptoms. This suggests that whereas lower overall segregation of the salience network is associated with more severe MBI symptoms, the functional disconnection specifically between the salience and default-mode networks may exacerbate MBI symptoms. The default-mode network has been associated with the integration of cognitive and emotional experience, as well as with emotion regulation.^25–27^ Thus, functional disconnection between the salience and default-mode networks may impair the regulatory influences of the default-mode network over emotional responses initiated by the salience network.

Our study has several limitations. First, the cross-sectional nature of our dataset makes it impossible to infer any causal relationships between tau positivity, salience network segregation, and MBI severity. Longitudinal studies are necessary to establish the temporal dynamics of these associations. Second, while PET biomarkers offer measurement of neuropathological burden, the resolution of PET imaging may not fully capture the microstructural changes impacting network functionality. Finally, while completely novel, our results should be considered preliminary due to the relatively small sample size; the present findings need to be replicated by future work.

In conclusion, we propose a novel model in which tau pathology influences MBI through alterations in the functional architecture of the salience network. By demonstrating that tau positivity is associated with reduced segregation of the salience network, and that such alterations are associated with increased severity of MBI symptoms, our findings suggest a critical role of network integrity in the pathogenesis of neuropsychiatric symptoms in AD. This work adds to the growing body of evidence supporting the concept of neurodegenerative diseases as network dysfunctions, where clinical manifestations are directly tied to the integrity of specific neural circuits. Moreover, our results offer a potential resolution to previous inconsistencies in the literature, suggesting that the functional status of the salience network may be a key factor in understanding the relationship between AD neuropathology and behavioral features. We encourage future research aimed at further delineating the interactions between neuropathological markers and neural network dynamics in AD and related disorders.

## Data Availability

The data sets analyzed for this study may be available upon reasonable request to the corresponding author, pending approval from participants and adherence with the NIH and local approval mechanisms.

## CONSENT STATEMENT

The University of Michigan Institutional Research Board approved this study. All participants or their legally authorized representative provided written informed consent (and assent as appropriate).

