## Supplement for "Salience Network Segregation Mediates the Effect of Tau Pathology on Mild Behavioral Impairment"

**Supplementary Figure**

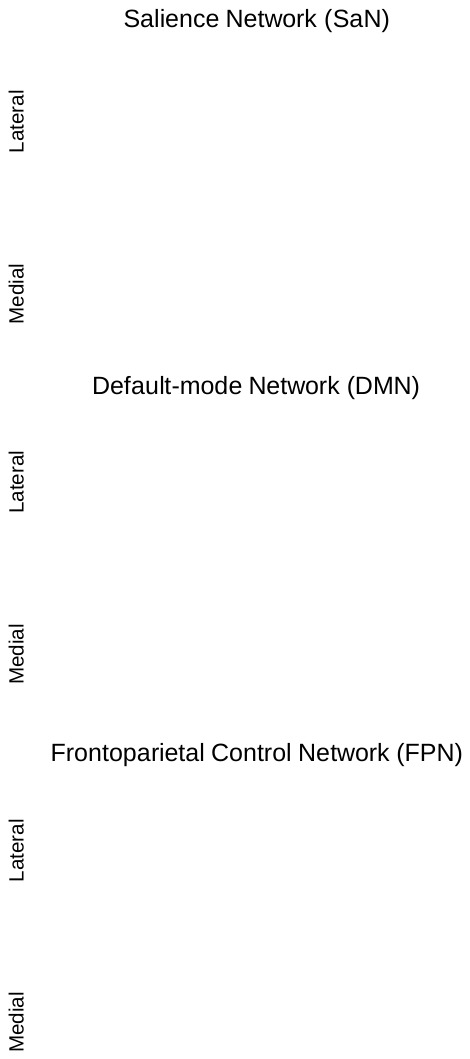

Supplementary Figure 1. Brain networks examined in the present study using the Power et al. (2011) atlas (see Methods section). Abbreviations: L: Left; R: Right.

**Supplementary Analysis**

We performed two regression-based analyses to test the paths proposed in our main model and determine whether the mediators (i.e., the three functional brain metrics: overall segregation of the salience network, overall segregation of the default-mode network, and pairwise salience–default-mode network segregation) carry the influence of the independent variable (AD biomarker status) to the dependent variable (MBI symptoms severity).^63^ This analysis was conducted using Stata^62^ statistical software.

First, a multivariate regression of the three functional brain metrics (overall segregation of the salience network, overall segregation of the default-mode network, and pairwise salience–default-mode network segregation) on AD biomarker status—accounting for effects of sex, age, education, cognitive performance (MoCA score), GM atrophy, percent outlier volumes, and inside-scanner motion—yielded a significant effect of tau-PET positivity on the overall segregation of the salience network (b=-0.08, p=0.004). This indicated that, relative to biomarker negative individuals, those who were both A*β* and tau positive showed lower segregation of the salience network from the other association networks.

Second, a negative binomial regression of MBI symptoms severity on the three functional brain metrics (i.e., overall segregation of the salience network, overall segregation of the default-mode network, and pairwise salience–default-mode network segregation) and AD biomarker status—accounting for effects of sex, age, education, cognitive performance (MoCA score), GM atrophy, and motion)—showed that lower overall segregation of the salience network was associated with greater severity of MBI symptoms (b=-3.95, p=0.023). In addition, greater pairwise salience–default-mode network segregation was associated with more severe MBI symptoms (b=5.72, p=0.003).

Taken together, these findings indicate a mediation effect of tau-PET positivity through the overall segregation of the salience network and no direct influence of AD biomarker status on the severity of MBI symptoms severity, consistent with the main results.
